# Shared Genetic Architecture Between Eating Disorders, Mental Health Conditions, and Cardiometabolic Diseases: A Comprehensive Population-Wide Study Across Two Countries

**DOI:** 10.1101/2024.10.20.24315825

**Authors:** Joeri Meijsen, Kejia Hu, Dang Wei, Stefana Aicoboaie, Helena L Davies, Ruyue Zhang, Mischa Lundberg, Richard Zetterberg, Joëlle Pasman, Weimin Ye, Thomas Werge, Cynthia M. Bulik, Fang Fang, Alfonso Buil, Nadia Micali

## Abstract

Eating disorders arise from a complex interaction of genetic and environmental influences. Here we provide comprehensive population-level estimates of the heritability of eating disorders and their genetic relationships with various mental health and cardiometabolic disorders (CMDs), expanding beyond genome-wide association studies. We examined the heritability of three eating disorders—anorexia nervosa (AN), bulimia nervosa (BN), and other eating disorders (OED)—and investigated shared familial and genetic risk factors with mental health disorders and CMDs. Using national-register data from Denmark and Sweden (1972-2016), we analyzed clinical diagnoses for over 67,000 individuals with eating disorders, their first-degree relatives, and matched controls from populations totaling 17 million. Heritability estimates were moderate, 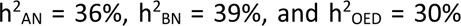 and genetic correlations revealed substantial overlap between AN and obsessive-compulsive disorder (r_g_ = 0.65) and moderate correlations with other mental health disorders such as autism (r_g_ = 0.36). Significant genetic associations were also identified between eating disorders and CMDs, showing strong replication across both countries. These findings emphasize the genetic foundations of eating disorders and their shared genetic architecture with mental health and CMDs. This research enhances our understanding of comorbidity patterns and has important implications for developing integrated treatment approaches.

## Introduction

Eating disorders (ED) such as anorexia nervosa (AN), bulimia nervosa (BN), binge-eating disorder (BED), and other eating disorders (OED) are often chronic and debilitating disorders occurring in 5-10% of the general population.^1^ As with all psychiatric disorders, both genes and the environment contribute to the risk of developing an ED. The role of genetics in eating disorders has been well-established using several study designs (e.g., family- and twin studies),^2^ and more recently via molecular techniques such as genome-wide association studies (GWAS).^3–5^ These different study designs have provided ample evidence of the heritability of eating disorders, the occurrence within families, the potential involvement of specific gene variants, and the genetic correlations with other disorders. Heritability estimates ascertained from twin studies vary across eating disorders. ^6^ For instance, of these three disorders AN has the highest reported twin-based heritability but also a broad range of estimates (0.28-0.74),^7–9^ followed by BN (0.55-0.62),^9–12^ and BED (0.39-0.45).^11,13,14^ Although not the focus of this study, a recent study has shown high twin-based heritability (0.79; 95% CI: 0.70-0.85) of broad avoidant restrictive food intake disorder (ARFID).^15^ Moderate heritability estimates for individual symptoms of eating disorders have also been shown, such as weight and shape concern (0.43), ^16,17^ binge eating (0.49),^16,17^ and self-induced vomiting (0.72),^18^ with some variability across studies.^19^

Historically, genetic studies, such as GWAS, have focused heavily on AN.^3,4,20–22^ However, a recent GWAS identified multiple common genetic risk variants associated with a BED phenotype obtained via machine-learning.^23^ The discovery of more risk variants is likely given that a larger AN GWAS and the first binge-eating GWAS are currently underway and nearing completion.^24^ Both twin studies and GWAS (specifically those of AN) have demonstrated the genetic overlap across eating disorders,^10^ and between eating- and other psychiatric disorders.^3,4,25,26^ The strongest positive genetic correlations with AN have been identified for obsessive-compulsive disorder (OCD), major depressive disorder (MDD), schizophrenia, and anxiety. In contrast, negative genetic correlations have been shown with cardiometabolic- and anthropometric traits (e.g., body fat percentage, insulin resistance, and leptin).^3,4^ In a recent study, we showed that whilst AN, BN, and BED share genetic risk with other psychiatric disorders, they differ in their shared genetic risk with cardiometabolic- and anthropometric traits.^5^ For example, whereas BED has positive genetic similarity with waist circumference and obesity, opposite patterns were observed for AN.

Twin studies and GWAS are useful in estimating the heritability and genomic architecture of complex traits and diseases; however, they have methodological limitations. First, both methods are often applied to highly self-selected clinical samples or rely on self-reported data, leading to a study sample that is not representative of the general population and thus potentially over- or under-estimating heritability. Second, single nucleotide polymorphism (SNP)-based heritability estimates are consistently lower than twin-based ones.^27^ This is in large part because GWAS exclusively assesses the association between common additive genetic variants and a disorder, and thus does not capture the effect of gene-environment interactions, rare variants, and other non-additive contributions. The greatest limitation of twin-based studies is reliance on the equal environment assumption, which assumes that mono- and dizygotic twins are similarly exposed to relevant environmental factors, biasing heritability estimates upwards. Indeed, a recent register-based study in Denmark supported the non-representativeness of twins as it showed that compared to singletons, twins carry a 40% increased risk, whilst triplets/quadruplets a 92% increased risk for developing AN.^28^ Therefore, the estimates using twin populations only may not reflect the genomic architecture of the general population, as they are likely to be biased upwards.

Building on existing knowledge and recent methodological advances,^29,30^ we leveraged nationwide healthcare registers and near-complete population genealogies of ∼17 million individuals spanning four generations across Denmark and Sweden, to: a) estimate the risk of eating disorders in first-degree relatives of probands with eating disorders; b) determine the heritability of three eating disorders; and c) investigate co-heritability (i.e., the genetic correlation) amongst specific eating disorders and, between eating disorders and multiple psychiatric disorders and cardiometabolic diseases.

## Methods

### Case definition

This study was based on the Danish and Swedish civil- and national health registries, which have been previously described in detail.^31–38^ Eating disorder cases included all individuals born in Denmark or Sweden who were clinically diagnosed with either AN, BN, and/or OED (Table S1) according to inpatient and outpatient discharge records from all Danish and Swedish hospitals up to December 31, 2016, obtained from the Danish and Swedish National Patient Register or Psychiatric Central Research Register (for details about years of coverage of both registers see).^38^ We applied the same criteria to identify cases for attention-deficit/hyperactivity disorder (ADHD), anxiety disorders, autism spectrum disorders (ASD), bipolar disorder, MDD, obsessive compulsive disorder (OCD), schizophrenia, type 2 diabetes, heart failure, hypertension, coronary artery disease, obesity, and peripheral artery disease cases. The total number of cases is less than the sum of all individual disorder diagnoses because many individuals have more than one diagnosis. Most AN, BN, OED, MDD, schizophrenia, and bipolar disorder diagnostic codes translate poorly to individuals below the age of 10, therefore we excluded individuals who received a diagnosis before this age. Family members (non-twin full-siblings and parents) were identified using the near-complete population genealogies derived from the multi-generation registers.^39^. Information about being male or female was obtained from the civil registers and for most individuals born in Denmark and Sweden refers to sex, which was assigned at birth. However, for some individuals this information should be referred to as gender as individuals are allowed to request a change to this information based on their preferred gender identity. Details regarding exclusion criteria and ICD-8, 9, and 10 codes used in this study are reported in Table S1.

### Statistical analysis

Primary analyses were cumulative incidence functions (CIF) with age at first hospital contact as an inpatient or outpatient for all eating disorders, other mental health disorders, and cardiometabolic diseases (CMD). CIFs were estimated for: a) the general population, and for individuals with any b) full sibling or c) parent diagnosed with the studied disorder, and finally for individuals with any d) full sibling or e) parent diagnosed with a diagnosis different than their own disorder (e.g., all offspring with AN with at least one parent diagnoses with MDD). All CIFs were calculated using the Nelson-Aalen estimator for individuals born in the same calendar year to account for substantial changes over time in the underlying incidence as well as censoring and competing risks.

We calculated the additive heritability (h^2^) under the liability threshold model based on the cumulative incidence as a function of pedigree relatedness. Briefly, we used the general population and full sibling CIFs at the last overlapping observed time point as estimates of the proportion of the population born in the same calendar year that were affected in their lifetime. Using these estimates, we calculated h^2^ estimates as described by Wray and Gottesman^29,40,41^ per given calendar year. All within-country h^2^ estimates were first meta-analysed using random effect inverse variance weighting followed by a between-country meta-analysis using the same method.

We calculated the genetic correlation (*r*_*g*_) between two traits by deriving the general population cumulative incidence-based risk for both disorders (e.g., AN and SCZ) and full siblings’ cross disorder familial risk (i.e., the cumulative incidence risk for AN when having a full sibling with SCZ). We extracted the last possible time point for each birth-year-stratified CIF and calculated the *r*_*g*_ by inputting the previously calculated h^2^ estimates using the formula described by Wray and Gottesman^29,40,41^. In line with the h^2^ calculations, all birth-year specific genetic correlation estimates were meta-analysed first within-country and followed by a between country meta-analysis. We repeated the analyses using parent-offspring comparisons. Danish register data was stored on a PostgreSQL 13.3 database server information was extracted using the psql 16.2 database client. All register-based analyses were done in R v4.2.1 using the cmprsk v2.2 package.

## Results

### Descriptive statistics

The prevalence of AN and BN was close to 0.5% in both countries but was higher in Denmark than in Sweden for individuals born between 1968-2000 (AN: 0.59% vs 0.53% and BN: 0.48% vs 0.32%). These differences were more pronounced within the younger (1985-2000) cohort (AN: 0.80% vs 0.72% and BN: 0.55% vs 0.37%). Conversely, the prevalence of OED in Sweden was higher than that of Denmark in both the older (1.02% vs 0.6%) and younger cohort (1.40% vs 0.79%). As expected, in both countries individuals who received an eating disorder diagnosis in were substantially more likely to be female than male (13:1). However, we observed small differences in sex-specific eating disorder prevalence between Denmark and Sweden; with BN having a higher prevalence in males in Sweden (3.4% versus 2.5%, *p*=1.19×10^-3^) and OED being observed more commonly in Danish males (9.3% versus 7.6%, *p*=1.01×10^-8^). We observed a significant difference in the use of in- and outpatient registration between the two countries (Table 1). For instance, Danish individuals with an AN or OED diagnosis were more likely to be treated exclusively as an inpatient compared to Swedish individuals with the same diagnosis (OR_AN_=3.83, OR_OED_=1.19) who were more likely to be treated via the outpatient hospital system (Table S2A). The opposite relationship was observed for BN. Overall, Danish individuals were significantly more likely to have both an in- and outpatient recording for all three EDs, potentially suggesting a hospitalisation trajectory. Individuals in Sweden were significantly younger when receiving their first AN diagnosis (median age in years: 17.8 versus 18.6, *p* =9.74×10^-11^) or OED (20 versus 21, *p*=7.09×10^-17^), whereas Danish individuals received a BN diagnosis at a younger age (24.3 versus 26.3, *p*=1.30×10^-47^). Across all eating disorder diagnoses, Danish individuals were, on average, born earlier than Swedish individuals and diagnosed earlier, based on calendar years (Table 1 and Figures S1-S3). These differences in prevalence, age at first diagnosis, year of birth, and year of diagnosis were too small to be clinically impactful but statistically significant due to the large sample size.

**Table 1.** Demographic characteristics by eating disorder in Denmark and Sweden. *The chi-square test was used for testing the difference in type of diagnosis and sex, two-sample z-test was used to assess the difference in prevalence, and Wilcoxon/Mann–Whitney U test was used for comparisons of continuous variables (year of birth, year of diagnosis, and age at first diagnosis) between Denmark and Sweden. Comparisons of continuous outcomes were performed in compliance with Danish and Swedish privacy laws*.

|  | Anorexia nervosa |  |  | Bulimia nervosa |  |  | Other eating disorder |  |  |
| --- | --- | --- | --- | --- | --- | --- | --- | --- | --- |
|  | Denmark | Sweden | *p* | Denmark | Sweden | *p* | Denmark | Sweden | *p* |
| **N** | 13,224 | 16,856 |  | 8,108 | 7,583 |  | 11,392 | 28,444 |  |
| Only outpatient | 2382 (18.0%) | 8728 (51.8%) | $<1 \times 10^{-99}$ | 6501 (80.2%) | 5888 (77.6%) | $1.09 \times 10^{-4}$ | 7871 (69.1%) | 21,624 (76%) | $4.8 \times 10^{-46}$ |
| Only inpatient | 6891 (52.1%) | 3729 (22.1%) | $<1 \times 10^{-99}$ | 291 (3.6%) | 740 (9.8%) | $1.47 \times 10^{-54}$ | 1534 (13.5%) | 3293 (1.6%) | $1.97 \times 10^{-7}$ |
| Both in and outpatient | 3951 (29.9%) | 4399 (26.1%) | $4.1 \times 10^{-13}$ | 1316 (16.2%) | 955 (12.6%) | $1.13 \times 10^{-10}$ | 1987 (17.4%) | 3527 (12.4%) | $1.65 \times 10^{-39}$ |
| **Male (%)** | 863 (6.52%) | 1054 (6.3%) | 0.34 | 202 (2.5%) | 256 (3.4%) | $1.19 \times 10^{-3}$ | 1064 (9.35%) | 2164 (7.6%) | $1.01 \times 10^{-8}$ |
| **Prevalence and SE (%)** |  |  |  |  |  |  |  |  |  |
| 1985-2000 | 0.8 ($1.23 \times 10^{-6}$) | 0.72 ($6.50 \times 10^{-9}$) | $<1 \times 10^{-99}$ | 0.55 ($1.21 \times 10^{-6}$) | 0.37 ($5.04 \times 10^{-7}$) | $<1 \times 10^{-99}$ | 0.79 ($1.46 \times 10^{-8}$) | 1.40 ($1.54 \times 10^{-8}$) | $<1 \times 10^{-99}$ |
| 1968-2000 | 0.59 ($3.97 \times 10^{-7}$) | 0.53 ($2.23 \times 10^{-9}$) | | 0.48 ($3.6 \times 10^{-7}$) | 0.32 ($2.03 \times 10^{-8}$) | | 0.6 ($5 \times 10^{-7}$) | 1.02 ($5.63 \times 10^{-9}$) | |
| **Year of birth**<br>*Mean, Median, SD* | 1981.5, 1985, 15.94 | 1985.2, 1988, 12.3 | $3.65 \times 10^{-60}$ | 1981.8, 1982, 9.79 | 1982.6, 1984, 10.0 | $5.43 \times 10^{-13}$ | 1982.3, 1986, 16.31 | 1985.6, 1989, 12.0 | $1.27 \times 10^{-16}$ |
| **Year of first diagnosis**<br>*Mean, Median, SD* | 2003.2, 2006, 11.1 | 2005.9, 2007, 7.9 | $3.30 \times 10^{-51}$ | 2006, 2007, 6.68 | 2008.9, 2009, 4.9 | $<1 \times 10^{-99}$ | 2007.7, 2010, 7.5 | 2008.8, 2010, 5.9 | $4.81 \times 10^{-24}$ |
| **Age at first diagnosis**<br>*Mean, Median, SD* | 21.7, 18.6, 10.3 | 20.7, 17.8, 9.0 | $9.74 \times 10^{-11}$ | 24.3, 22.6, 7.57 | 26.3, 23.9, 8.8 | $1.30 \times 10^{-47}$ | 25.4, 21, 14.1 | 23.2, 20, 10.1 | $7.09 \times 10^{-17}$ |

We observed a statistically significant difference between the number of eating disorder diagnoses per individual in each country. For instance, individuals diagnosed in Denmark were substantially more likely to have an exclusive diagnosis of AN (OR=1.88, *p*<1×10^-99^) or BN (OR=2.54, *p*<1×10^-99^), whilst individuals diagnosed with an eating disorder in Sweden were more likely to have exclusively an OED diagnosis (OR=0.41, *p*<1×10^-99^). If multiple eating disorder diagnoses were given, Swedish individuals were more likely than Danish individuals to receive a diagnosis of AN (OR=0.65, *p*=4.74×10^-75^) or BN (OR=0.84, *p*=4.27×10^-7^) together with an OED diagnosis (Figure 1), whereas Danish individuals were more likely to have both a diagnosis of AN and BN (OR=3.06, *p*=1.22×10^-83^) compared with individuals in Sweden. More can be found in Supplementary Table S2A-B.

**Figure 1:**
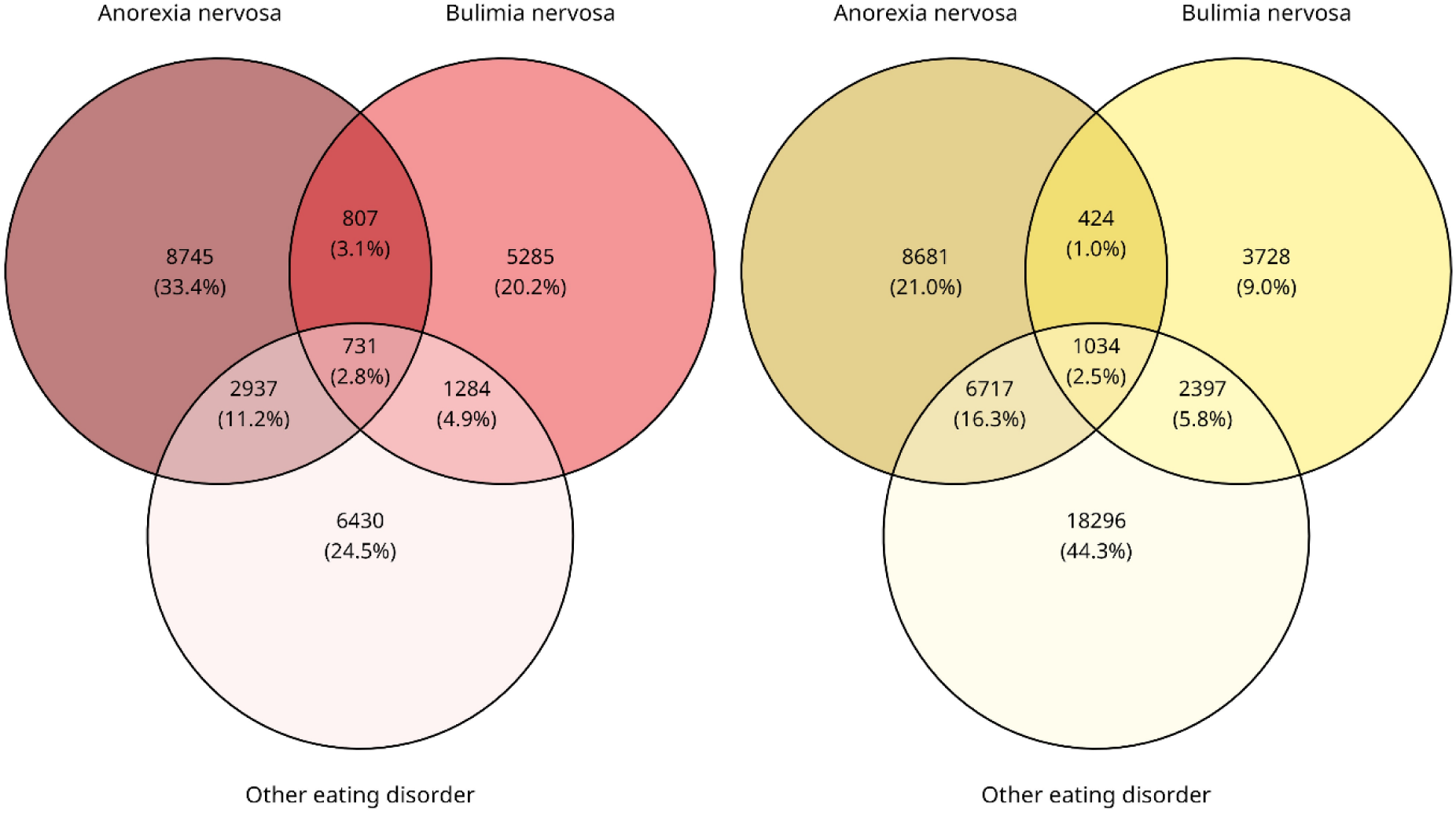
Venn diagram of eating disorder diagnoses made in left.) Denmark (n=26,219) and right.) Sweden (n=41,277). *Percentages reported are calculated as the number of individuals in each diagram section divided by the total number of individuals diagnosed in each country.*

### Familial risk

To assess the familial risk of eating disorders, we compared CIFs between individuals with a first-degree family member (i.e., parents and siblings) diagnosed with an eating disorder relative to the risk in the general population. The incidences were calculated from the number of new cases occurring for each age year. We calculated cumulative incidences up to a maximum of age 49 for birth cohort 1968-2000 and age 32 for birth cohort 1985-2000. We observed that individuals were substantially more likely to receive an eating disorder diagnosis if they had a first-degree relative (i.e., parent or full sibling) diagnosed with the same disorder (Figure 2). In general, the probability of an eating disorder increased on average three-fold for all disorders when having either a parent or full sibling with an eating disorder diagnosis. However, in Sweden, children were less likely to receive a BN diagnosis if they had a parent with a BN diagnosis, regardless of birth cohort, compared to Danish individuals who had an increased risk of 1.5 to 8.3-fold. This might, in part, be due to differences in recording time and left-sided censoring of medical data, as well as children of BN-diagnosed individuals in Sweden being too young to be diagnosed themselves. All estimates can be found in Table S3.

**Figure 2:**
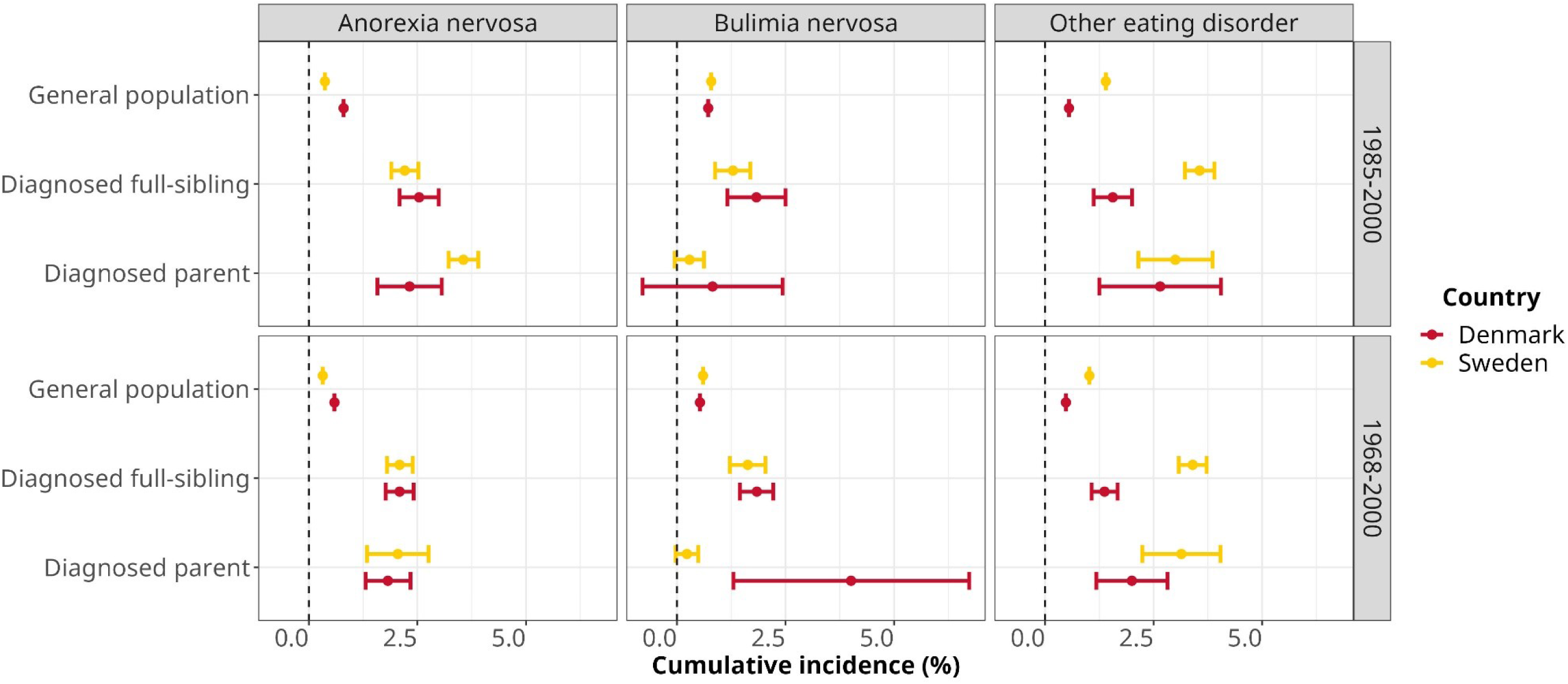
Familial risk of eating disorders in Denmark and Sweden. *Cumulative incidences (%) and 95% confidence intervals for eating disorders were calculated for different birth cohorts (1985-2000 and 1968-2000) using medical records up to 2016 including: the general population (all individuals born in birth window), individuals with at least one full sibling diagnosed with the same eating disorder, and individuals with at least one parent diagnosed with the same eating disorder. Individuals diagnosed before age 10 were excluded. Risk estimates were taken, if possible, at age 49 (1968-2000) and 32 (1985-2000).*

### Heritability estimates (h^2^)

To estimate the heritability of eating disorders, we combined familial risk estimates as described in the methods section. We calculated 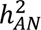 as 0.36 (95% CI 0.30-0.41), 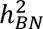 as 0.39 (95% CI 0.32-0.46), and 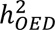 as 0.30 (95% CI 0.20-0.40, Figure 3). The h^2^ of AN and BN were broadly in line with previously reported estimates^2^ whereas the heritability of OED has, to our knowledge, not been studied enough to make an adequate comparison. We observed no statistically significant difference between country-specific estimates (Bonferroni *p*<3.33×10^-3^) (Table S4).

**Figure 3:**
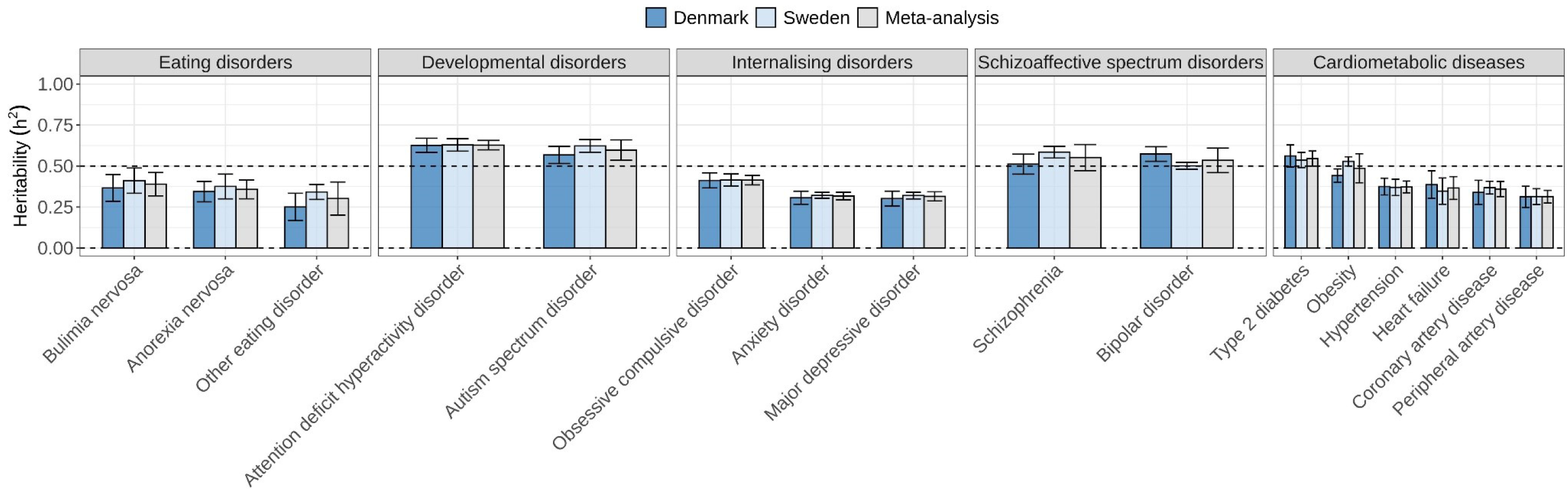
Narrow sense heritability (h^2^) estimates and 95% confidence intervals of eating disorders, other mental disorders, and cardiometabolic diseases in Denmark and Sweden using full-sibling comparisons. *All within-country (Denmark and Sweden)* ℎ^2^ *estimates reported per birth year were first meta-analysed using random effect inverse variance weighting followed by a between-country meta-analysis using the same method*.

Due to the large sex difference in eating disorder prevalence (>90% female) and small sample size in the Danish cohort, the heritability of eating disorders in females was near identical to the non-sex-stratified heritability. In comparison, the male estimates presented substantially large confidence intervals, or no heritability could be calculated (BN). We did not observe significant differences in sex-stratified h^2^ estimates of eating disorders in the Swedish sample (Table S4). Estimates of h^2^ of other mental health disorders and cardiometabolic diseases as well as all meta-analysed estimates of h^2^ including SE, CIs, and p-values are reported in Table S4.

We performed a sensitivity analysis to assess the effect of exclusion criteria on the heritability (Figure S4). First, we assessed the effect of in-patient and outpatient recording by estimating the h^2^ separately. The 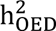 using inpatient ICD-10 diagnoses was 0.59 (95% CI 0.42-0.77), significantly higher (Bonferroni *p*<1.15×10^-3^) than the 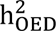 using outpatient ICD-10 diagnoses: 0.27 (95% CI 0.19-0.36) using Danish individuals and could not be replicated using Swedish data. No significant differences between in- and outpatient heritability estimates of AN and BN were observed. Furthermore, no significant difference was observed between the 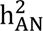 diagnosed under ICD-8 or ICD-9 (exclusively inpatient) and the 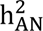 diagnosed as inpatient using ICD-10 (*p*=0.26). All estimates of h^2^ using various exclusion criteria can be found at Supplementary Table S5.

### Genetic correlations (r_g_)

We first quantified the genetic relationship across the three eating disorders via genetic correlation analyses. Next, we calculated genetic correlations across the three eating disorders by and performing multiple between disorder comparisons using a) seven other mental disorders, and c) six CMDs. All analyses were conducted using data from Denmark and Sweden using both full-sibling and parent-offspring comparisons. All estimates of *r*_*g*_ are reported in Table S6 and S7

#### Between eating disorders

Genetic correlation analysis between eating disorders using full-sibling information showed a large shared genetic contribution between eating disorders ranging from *r*_*g*_=0.69 to *r*_*g*_=0.96 (Table S6). Meta-analyses of Danish and Swedish estimates did not significantly change the estimates. The analysis was repeated by stratifying individuals by in-patient and outpatient eating disorder hospitalisation; we observed no significant change in genetic correlation estimates.

#### Eating disorders and other mental health disorders

We estimated genetic correlations for each of the 21-eating disorder and other mental disorder diagnostic pairs using full-sibling comparisons and observed, again, similar estimates between countries (Figure 4). After meta-analysis, 25 genetic correlations were statistically significant (Bonferroni *p*<1.39×10^-3^) (Figure 4). Genetic correlations for all the ED-other mental disorder pairs were positive and statistically significant except AN-schizophrenia. In line with previous literature^42^, OCD showed the largest genetic correlation with both AN (*r*_*g*_=0.65, 95% CI 0.56-0.73) and OED (*r*_*g*_=0.66, 95% CI 0.53-0.79). Other large genetic correlations were observed between OED and other mental disorders such as MDD (*r*_*g*_=0.57, *p*=1.01×10^-131^), ADHD (*r*_*g*_=0.43, *p*=2.03×10^-96^), anxiety disorders (*r*_*g*_=0.57, *p*=1.59×10^-79^), and ASD (*r*_*g*_=0.46, *p*=6.30×10^-65^). Genetic correlations between the eating disorders and ADHD and ASD were not replicated using parent-offspring comparisons. Overall, estimates from Denmark replicated well with Swedish estimates, with one genetic correlation showing a significant difference between countries using full-sibling information and five using parent-offspring data. In general, our results suggest that the genetic architecture of eating disorder is widely shared (between 0.14 and 0.69) with other mental disorders.

**Figure 4:**
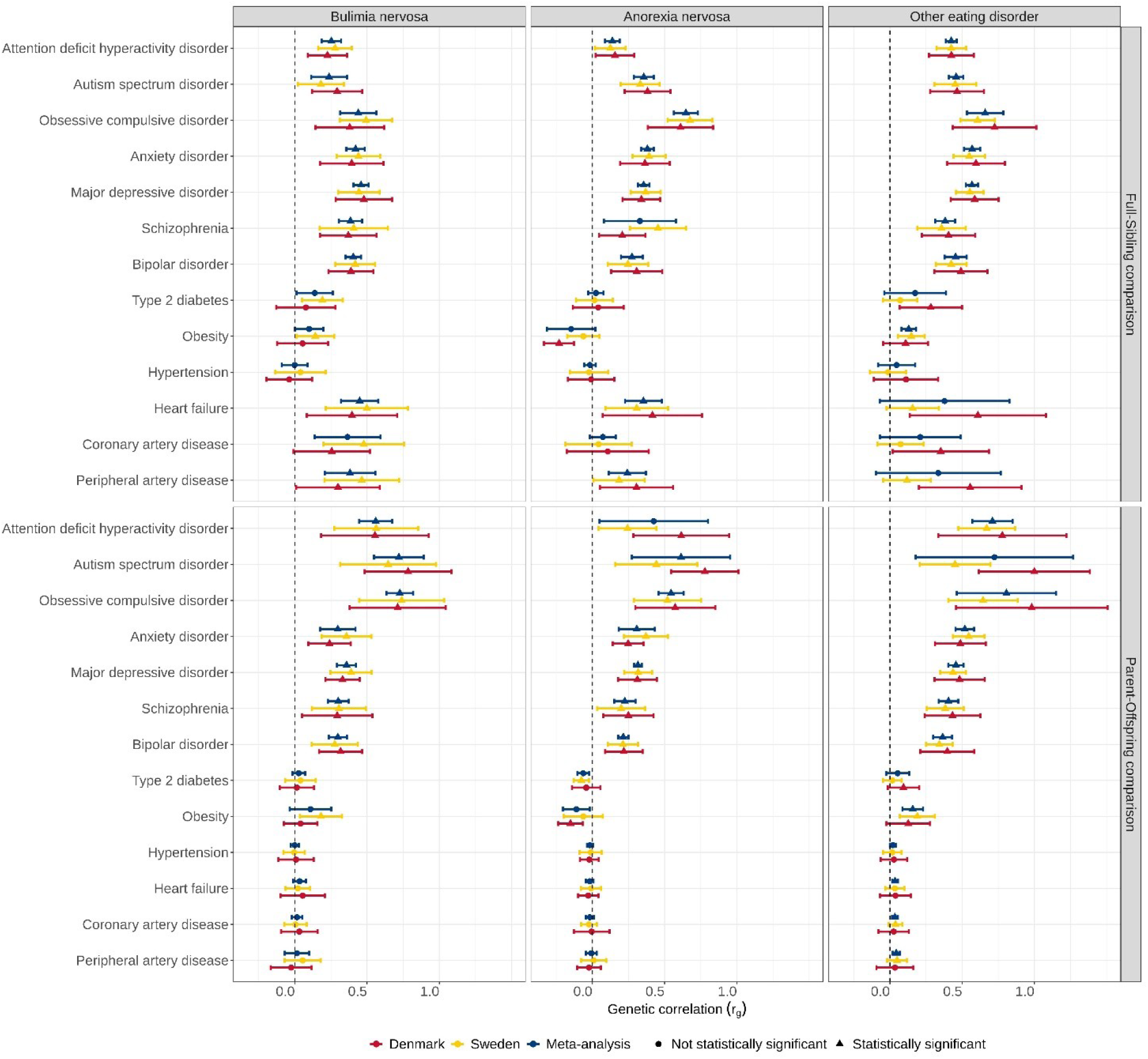
Danish and Swedish genetic correlations (r_g_) estimates and 95% confidence intervals between eating- and other mental disorders, and between eating- and cardiometabolic disorders calculated using national register data. *All within-country (Denmark and Sweden) r*_*g*_ *estimates reported per birth year were first meta-analysed using random effect inverse variance weighting followed by a between-country meta-analysis using the same method. Top row panels r_g_ estimates based on full-sibling comparison, bottom row panels r_g_ based on parent-offspring comparisons. Bonferroni p<1.28×10^-3^*

#### Eating disorders and cardiometabolic diseases

We observed significant positive genetic correlations between AN, BN and two CMDs (heart failure, and peripheral artery disease) in full-sibling comparisons (Figure 4) and five estimates were significantly different between Denmark and Sweden. The observed significant genetic correlations between AN, BN and two CMDs did not replicate using parent-offspring comparisons Finally, no significant associations were observed between obesity and AN and BN. However, obesity shares a substantial genetic overlap with OED (*r*_*g*_=0.13, *p*=5.46×10^-7^) which was replicated using parent-offspring comparisons (*r*_*g*_=0.16, *p*=1.71×10^-5^). All but one *r*_*g*_ (OED and obesity) using parental information were similar between countries.

As sex stratification did not seem to affect the heritability, or no (reliable) heritability could be estimated using the Danish data, we opted not to perform any sex stratified genetic correlation analysis.

## Discussion

Here we have shown that the h^2^ estimates were substantial i.e., 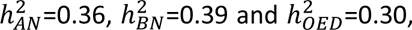 and not significantly different across eating disorders. Reducing phenotypic heterogeneity by excluding individuals with prior eating disorder diagnoses other than the eating disorder of interest did not affect the h^2^. The h^2^ of, ICD-10 OED in Denmark for individuals receiving inpatient care was two times larger (0.60) than the h^2^ of ICD-10 OED diagnosed in outpatient care (0.27), suggesting stronger genetic underpinnings for eating disorders with greater severity.^43^

The observed pattern of genetic correlations with mental disorders and cardiometabolic diseases advances understanding of the underlying aetiology, comorbidities, and complexity of different eating disorders. The largest positive genetic correlation observed between AN and OCD (*r*_*g*_=0.65, *p*=6.68×10^-48^) is in line with previous family,^48,49^ register,^26^ and genetic studies.^42^ Moreover, previous analyses grouped mental disorders based on shared genomics and clustered AN, OCD, and Tourette syndrome into one category.^50^ These findings therefore point towards a higher-level genetic structure that underlies disorders characterized by compulsivity^42^ compared to other mental disorders. In addition, eating disorders are often comorbid with internalising disorders, e.g. multiple anxiety phenotypes^51,52^ and mood disorders,^53,54^ which was also observed in this study.

The positive genetic correlations seen in this study between AN and ASD (*r*_*g*_=0.36, *p*=2.50×10^-22^) support the increasing hypothesis of a substantial genetic overlap between these disorders.^55^ Several clinical studies have hinted an overlap between eating disorders (predominantly AN) and ASD, given the overlapping features such as limited emotional expression, reduced social contacts, and cognitive rigidity,^56,57^ yet no significant genetic associations have been observed previously.^4,58^ Differences in full-sibling and parent-offspring genetic correlations are likely due to increased awareness and diagnosis of neurodevelopmental disorders.

In line with recent work^5^, we did not observe a significant negative genetic correlation between AN and type- 2 diabetes using parent-offspring comparisons (*r*_*g*_=-0.06, *p*=2.50×10^-3^) and when using sibling comparisons (*r*_*g*_=0.02, *p*=0.37). However, this warrants further investigation as other large GWAS using linkage-disequilibrium score regression have observed significant negative genetic correlations between the two outcomes.^3,4^ Nor did we observe a significant negative genetic correlation between AN and obesity (*r*_*g*_=-0.11, *p*=0.02)^4^; however this is likely due to the nature of our sample, as obesity is likely to be diagnosed in the registers only if relevant to pathological findings in the healthcare context, and will be under-represented in this context. The discrepancy between previously observed negative genetic correlations between AN and type-2 diabetes can partially be explained by differences in sampling of individuals across different types of studies. In general, GWAS are performed using non-population representative samples whereas a national register-based study contains all individuals with diagnoses that have been in contact with the hospital at any time and therefore mostly free from generally occurring biases (e.g., volunteering bias and selection bias). In this case it is entirely plausible that observed negative genetic correlation using GWAS data may in fact be influenced by other hidden and co-occurring disorders. Our finding of a relationship between AN, BN and two CMDs (heart failure and peripheral artery disease) using (young) full-sibling comparisons is novel and warrants further investigation. The large overlap between OED and obesity is potentially hinting at an enrichment of binge-eating disorder diagnoses in OED, which is not defined as a separate diagnostic entity in ICD-10. Our findings overall highlight the need to implement current diagnostic manuals (ICD-11) in research that do include BED as a separate diagnosis, given the impact this has on a large group of the population. In contrast to the genetic correlations calculated between EDs and other disorders, the genetic correlations between the eating disorders were substantially larger and often not significantly different from one, suggesting a high degree of genetic similarity across all EDs. Stratification based on hospital admission type (i.e., in or outpatient) did not affect the genetic correlation estimates, which may rule out disorder severity affecting these genetic correlations.

Although the genetic correlations were comparable between the two familial relationships in our study, we observed substantially larger confidence intervals between specific clusters of disorder constellations related to differences in age of onset. For example, most mental disorders have a relatively early age of onset whereas CMDs tend to be diagnosed later in life. This, in part, explains why the confidence intervals of the genetic correlations between eating disorders and CMDs are substantially smaller using parent-offspring information compared to full-sibling information as parents generally have lived long enough to develop a CMD. The opposite pattern was observed for eating disorders and other mental disorders, apart from schizophrenia, which has an older average at onset than other mental disorders.

Our study has some limitations. First, we utilised secondary care hospital diagnoses from 1960s onwards since the establishment of the inpatient registers in both countries. Therefore, some relatively newer diagnoses (e.g., BN) will be under-ascertained in older individuals and may skew the age at onset estimations. This underrepresentation might also result in misclassification (e.g., in Denmark) due to the non-adoption of ICD-9 and therefore, the “late” introduction of BN as a substantive diagnostic category. Second, the outpatient diagnoses were recorded decades later than the inpatient diagnoses in both countries, and diagnostic practices might have varied over time and between countries. However, we hypothesise that our calendar year-stratified analyses partially account for these time trends within and between countries. Another limitation concerns undiagnosed individuals, for instance, only ∼20-30% of individuals with an eating disorder in the general population seek or access medical services.^59–61^ Therefore, our cases likely represent more severely affected individuals who sought treatment and therefore is not be fully representative of the entire population of individuals affected by eating disorder, which in turn may have inflated our h^2^ and r_g_ estimates. Due to diagnostic manuals used in the healthcare systems in Denmark and Sweden, we were not able to differentiate other eating disorders outside of AN and BN, therefore our OED category is likely to be a heterogeneous group and might comprise multiple eating disorders that are undefined up to ICD-10 (e.g., BED).

Nonetheless, our study has several important implications for research and clinical practice. By quantifying heritability in a non-twin near-complete total population sample of two countries, we provide a more comprehensive estimate of the contribution of genetic factors to the variance of eating disorders. Second, we show an increased familial risk of eating disorders, namely that there is on average a three-fold higher risk among individuals who have a first degree relative with an ED compared to the general population. Quantification of familial risk can help prevention and early identification. Although the intergenerational transmission of eating disorders has been studied less than other mental disorders, ongoing studies will help understand risk markers and developmental trajectories. The identified genetic correlations between eating disorders and other somatic diseases and psychiatric disorders have supported both a deeper investigation of the pathophysiology of disorders,^5^ and a reconceptualization of eating disorders as disorders encompassing brain and body.^62^ This study confirms the strong genetic relationship between eating disorders and other psychiatric disorders, particularly internalizing disorders (e.g., OCD, anxiety, and depressive disorders). Clinically these disorders are highly comorbid with eating disorders and clarifying the nature of this overlap can improve clinical management and therapeutic approaches to comorbidities. For example, the overlap between ASD and eating disorders in clinical samples has received much attention recently,^63^ leading to the development of specific care pathways, and this paper is the first to provide a likely genetic basis for this observed overlap.^64,65^

In summary, we derived population-wide genetic estimates by harnessing the power of data containing almost all hospital records and genealogical information of two entire Scandinavian countries (*n*=17 million). Our results provide a unique perspective on the complex heritable nature of eating disorders as well as the overlap in genetic aetiology underlying psychiatric- and cardiometabolic diseases.

## Supporting information

All tables

## Ethical approval

The use of Danish data was approved by the Danish Health Data Authority (project no. FSEID-00003339) and the Danish Data Protection Agency. By Danish law, informed consent is not required for register-based studies. The use of Swedish data was approved by the regional ethics review board in Stockholm, Sweden with DNR 2012/1814-31/4.

## Data sharing statement

This work is based on Danish register data that are not publicly available due to privacy protection, including the General Data Protection Regulation (GDRP). Only Danish research environments are granted authorisation. Foreign researchers can, however, get access to data under Danish research environment authorisation. Further information on data access can be found at https://www.dst.dk/en/TilSalg/Forskningsservice or by contacting the corresponding authors. The use of Swedish data was approved by the regional ethics review board in Stockholm, Sweden with DNR 2012/1814- 31/4. Data from Swedish registers are not available for sharing due to policies and regulations in Sweden. Swedish register data are available to all researchers through applications at Statistics Sweden (SCB, https://www.scb.se/en/) and The National Board of Health and Welfare (Socialstyrelsen, https://www.socialstyrelsen.se/). By both Danish and Swedish law, individual consent to use register data for register-based studies is not required.

## Acknowledgement.

JM, AB, TW, and FF received funding from the European Union’s Horizon 2020 Research and Innovation Programme: the "predicting comorbid cardiovascular disease in individuals with mental disorder by decoding disease mechanisms" project (CoMorMent, grant number 847776, to Ole Andreassen). JM, JP, TW, and AB were supported by the US National Institutes of Health study on extreme MDD (R01 MH123724, to Patrick Sullivan). JM, NM, SA, and HD were supported by the Laureate Grant Award from the Novo Nordisk Foundation (Grant No: NNF22OC0071010, to NM). RZ was supported by the Swedish Research Council (Vetenskapsrådet, grant no. 2022-00242); JP received funding from the European Research Council (grant number 101042183). CB was supported by the National Institute of Mental Health (R56MH129437; R01MH120170; R01MH124871; R01MH119084; R01MH118278; R01 MH124871); Swedish Research Council (Vetenskapsrådet, award: 538-2013-8864); and Lundbeck Foundation (Grant no. R276-2018-4581).

## Author contribution

JM, AB, and NM conceived the study. JM, AB, CB, RZ, and NM contributed to the study design. JM, SA and HLD, performed the literature search and JM KH, DW, ML, JP, and RZe performed programming and/or data analyses. JM, AB, RZ, NM, FF, and TW contributed to data interpretation. FF, WY, AB, and TW provided access to data. JM, CB, and NM wrote the initial draft. NM, FF, and TW obtained the funding.

